# Understanding the Psychological Factors that Impact Hypertension: A Systematic Review

**DOI:** 10.1101/2025.08.21.25334144

**Authors:** Awele Rosemary Ebigwei Omeda, Marcus Chilaka, Masoud Mohammadnezhad, Eleftheria Vaportzis

**Author notes:** **Corresponding Author:** Eleftheria Vaportzis, School of Social Sciences, University of Bradford, Bradford, UK.

## Abstract

The development and management of hypertension strongly depends on psychological elements which include depression, anxiety together with stress and psychosocial support. This review analysed psychological elements that affect hypertension development from the year 2014 to 2025. The research identified 106 studies through a systematic database search of MEDLINE, Embase, PsycINFO, Scopus and CINAHL. Due to diverse methodologies, the research employed a narrative synthesis approach. The risk of bias was assessed using the Cochrane Risk of Bias Tool, Newcastle–Ottawa Scale, Domain-Based Approach, CASP, and ROB-MR Instrument. Results indicated that both depression and anxiety increased the risk of developing hypertension and decreased adherence to treatment. In contrast, mindfulness-based interventions showed potential blood pressure lowering effects, although evidence for long term outcomes is limited. People with strong psychosocial support networks and higher levels of life satisfaction had better medication adherence and lower stress levels. However, the current evidence base shows that most studies originate from high-income countries, with low- and middle-income countries having limited evidence and most low-income settings contributing only one or two studies, with no more than four studies per country. The results support the need to integrate physical and mental health care models in the management of hypertension. To enhance understanding of the psychological aspects of hypertension, future research should include underrepresented regions and implement both longitudinal and qualitative methods.

## Introduction

According to the World Health Organisation (WHO), hypertension, otherwise known as high blood pressure, affects 1.28 billion adults aged 30 to 79, with most cases found in low- and middle-income countries (LMICs) (WHO, 2023a). In clinical practice, medical professionals use pharmaceutical interventions together with lifestyle modifications to treat hypertension (WHO, 2023b). However, the development of hypertension represents a complex condition which goes beyond biological factors and lifestyle choices that require more than medication and lifestyle changes for effective treatment (Cuevas et al., 2017). According to Spruill (2010), hypertension emerges from the combination of biological elements with psychological environments that people experience across their lifespan.

Specifically, research outlines that psychological factors create complex pathways which affect cardiovascular health (McEwen, 2017). These pathways can be understood through: (1) the biopsychosocial model which explains these pathways through its emphasis on biological psychological and social factors that determine health outcomes (Engel, 1977). This model demonstrates that biological factors alone cannot explain health because psychological and social elements including stress, coping strategies and social networks affect biological processes and health results. (2) the concept of allostatic load by McEwen and Stellar (1993) describes how persistent psychological stress leads to progressive physical deterioration of the body. The body’s ability to maintain homeostasis becomes impaired while the cardiovascular system faces increased strain due to prolonged wear and tear which eventually leads to hypertension and other health issues.

Although most findings demonstrate psychological elements lead to higher blood pressure (BP), some studies present opposite findings which support the emotional dampening hypothesis which propose extended stress periods can produce diminished physiological reactions which might result in decreased BP readings (McCubbin et al., 2011). Despite these varying results and studies that have investigated the psychological factors affecting hypertension, the evidence base remains dispersed when considering different populations, study methodologies and designs. Past studies have investigated primarily single stressors such as anxiety, depressive states, stress, and PTSD but failed to combine findings about cognitive-behavioural responses alongside emotional regulation strategies and psychosocial interventions that interact with hypertension. In addition, there is a lack of culturally appropriate research in LMICs, even though hypertension and psychological stress commonly affect these regions (Londoño Agudelo et al., 2021; Schutte et al., 2021).

The purpose of this systematic review is to collate existing research evidence on how psychological factors affect hypertension development and management. It evaluates the connection between multiple psychological elements that include common mental health disorders, chronic stress, psychosocial interventions, socioeconomic factors and vulnerabilities of marginalized groups in hypertension management to understand their impact on hypertension. By compiling multiple recurring patterns from current research, the review spotlights understudied elements like regional characteristics in LMICs and qualitative data while also addressing essential knowledge gaps including the integration of cognitive-behavioural responses, and psychosocial interventions in hypertension management. This approach aims to create practical guidance for unified healthcare approaches.

## Methods

### Search Strategy

The research followed PRISMA 2020 guidelines for a systematic review. The protocol was prospectively registered on PROSPERO (ID: CRD42024531112). A search was performed using six databases including MEDLINE, Embase, PsycINFO, Scopus, PubMed, and CINAHL for studies from 1^st^ January 2014 to 1^st^ July 2025 that investigated psychological factors related to hypertension. The search strategy used Boolean operators and truncation techniques to combine keywords and MeSH terms including “hypertension,” “anxiety,” “stress,” “psychological disorders” for a broad yet targeted search. The full search strategy and references of the articles that met the inclusion criteria are presented in the supplementary materials.

### Eligibility Criteria

Studies were included if they contained human participants of any age, sex, or location and examined psychological factors related to the development of hypertension (e.g., anxiety, depression) or interventions for the management of hypertension (such as cognitive behaviour therapy, mindfulness). Eligible designs included quantitative (randomised controlled trials (RCTs), cohort studies, case-control studies, Cross-sectional), qualitative, and mixed-methods studies. and mixed-methods studies.

Primary outcomes were the incidence or management of hypertension. Secondary outcomes were changes in hypertension, psychological well-being, and intervention effectiveness. Only English-language studies published in peer-reviewed journals from 2014 were included. Studies were excluded if they were published in languages other than English, if they focused on comorbid conditions (such as diabetes, kidney disease, pregnancy), or if they focused on biomedical, lifestyle factors without addressing psychological aspects. Non-peer-reviewed articles, systematic reviews, case reports, case series, editorials, commentaries, or conference abstracts were also omitted.

### Study Selection

All records were imported into Covidence for deduplication and screening. From the 3,054 retrieved records, 1,499 unique studies remained after eliminating duplicates. The screening process was divided into two stages, which included title and abstract screening followed by full-text review. The first author (AREO) screened all articles. A second reviewer (EV) independently assessed 10% of the sample to ensure consistency. Discrepancies were resolved through consensus with a third reviewer (MC). Eighty-five studies met all inclusion criteria and were included in the final synthesis (see Figure 1).

**Figure 1:**
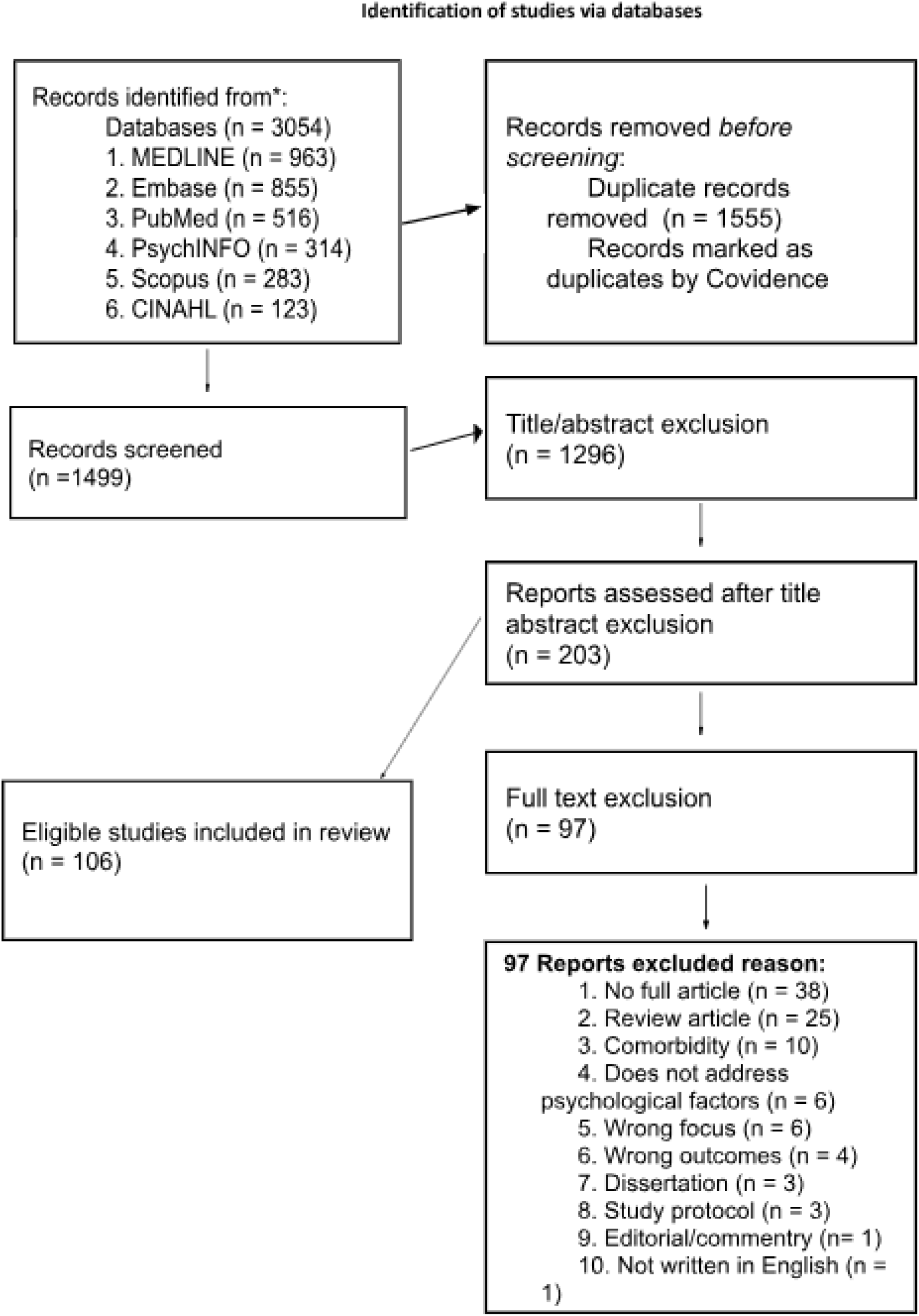
PRISMA flow diagram

### Data Extraction

Extracted information included study design, sample size, population characteristics (e.g., age range), psychological and hypertension outcomes, and study location. These research parameters were then synthesised to create an overview of the study characteristics, focus areas and geographical distribution of the included literature.

### Risk of Bias Assessment

The Cochrane Risk of Bias Tool was used to assess bias in RCTs across randomization, blinding and outcome reporting domains (Higgins et al., 2022). The Newcastle–Ottawa Scale (NOS) was used to assess cohort and case-control studies by evaluating selection, comparability and outcome/exposure assessment (Wells et al., 2014). An adapted NOS was used for Cross-sectional studies to fit the design’s methodological characteristics (Blanchard et al., 2024). The ROB-MR tool was used to evaluate Mendelian randomization studies by addressing bias sources that are specific to genetic instrumental variable analysis (Spiga et al., 2023). Pre-post and lab-based studies were assessed using a domain-based approach following Cochrane (Higgins et al., 2024) and NICE (2022) recommendations which focused on internal validity and measurement consistency. The CASP Qualitative Research Checklist was used to assess the trustworthiness and relevance of qualitative studies, focusing on research aims, methodology, ethical considerations, data analysis, and the value of the findings (CASP, 2025).

Of the 106 included studies, 18 were high quality, 86 moderates, and 2 exhibited notable methodological concerns.

### Data Synthesis and Analysis

A structured, theme-based synthesis and analysis was conducted to capture the diverse psychological influences on hypertension (Braun & Clarke, 2006). Thematic clusters were developed during full-text review, based on the included studies all of which employed quantitative methods. One qualitative study met the inclusion criteria, highlighting a gap in understanding lived experiences and context-specific psychological processes that are related to the development of hypertension.

This review examined different psychological factors (depression, anxiety, stress, and PTSD) in relation to hypertension through their strength of association which included strong, moderate, null, or inverse relationships. The thematic synthesis presented provides evidence to create practical recommendations which enhance hypertension care through psychological and social and contextual elements in treatment strategies.

## Results

### Study Characteristics and Geographic Scope

A total of 106 studies were eligible for this review, consisting of 61 Cross-sectional, 29 cohort, 6 RCTs, 3 case-control, 2 laboratory-based, 2 pre-post intervention, 2 Mendelian Randomization study, and 1 qualitative. These studies were conducted in 41 different countries. The United States (n = 27), China (n = 13), and the United Kingdom (n = 8) were the most represented countries among them (see Figure 2 Geographical Distribution), France, Ghana, the Netherlands, Colombia, Afghanistan, Peru, South Africa, Taiwan, Italy, and India. Ademola et al. (2019) included data from Nigeria and Ghana and Stein et al. (2014) included data from 19 countries.

**Figure 2:**
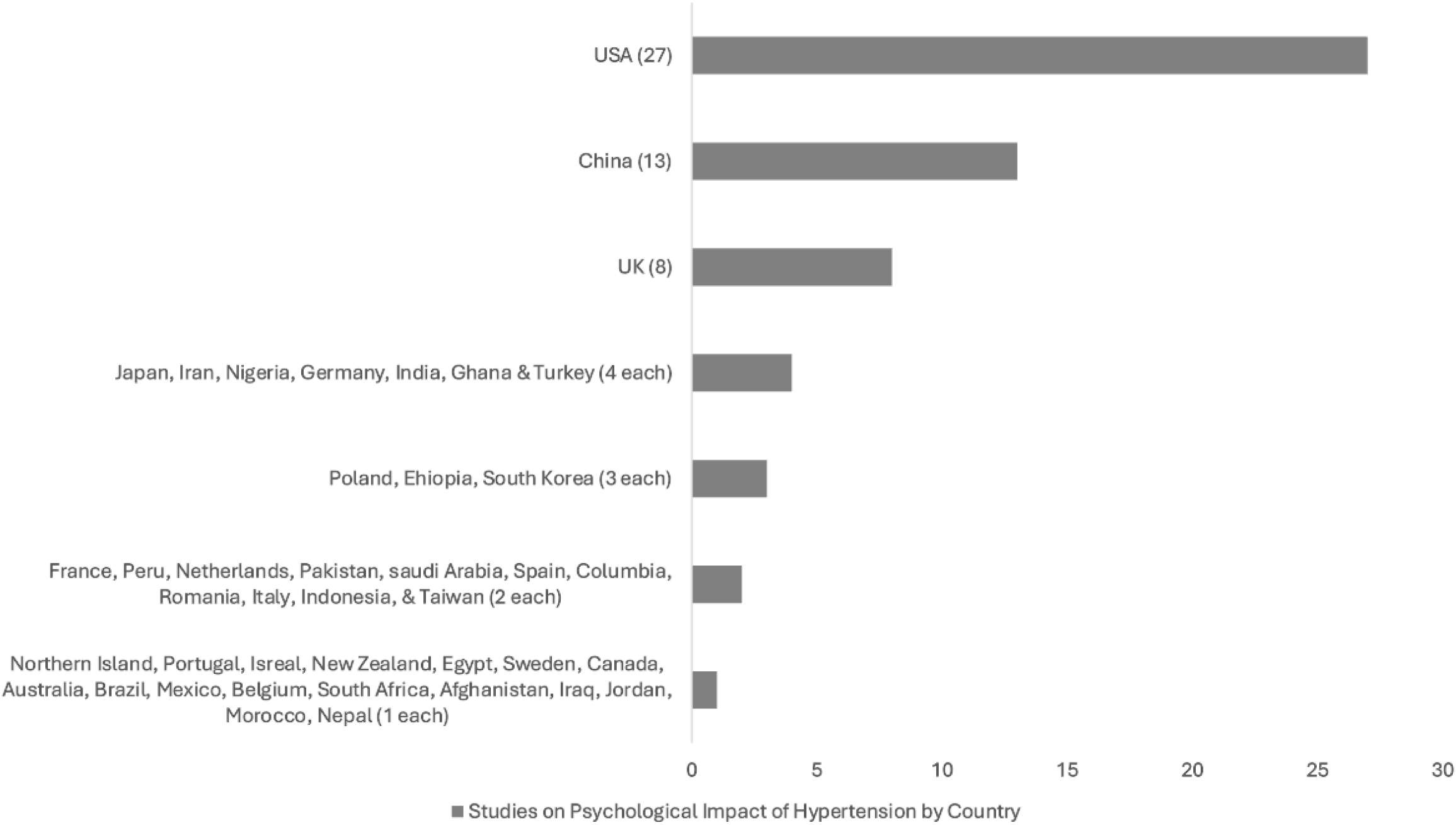
Showing the geographical distribution

The existing evidence base demonstrates that high-income countries produce most studies on psychological impact of hypertension, while LMICs lack sufficient representation because they contribute only one to two studies and no more than four studies each. Research capacity and infrastructure differences between regions may explain this geographic disparity especially in underrepresented areas such as sub-Saharan Africa and Latin America (Mills et al., 2020). As a result, the underrepresentation of certain populations hinders understanding of local stressors, cultural coping mechanisms, and healthcare disparities (Elnaem et al., 2022; Schutte et al., 2021).

### Age Distribution and Relevance to Hypertension Research

The research studies in this review examined participants across various age groups starting from adolescents through to older adults. Three studies included participants younger than 18 years old (Astudillo et al., 2024; Mucci et al., 2016; Santoni et al., 2025) which aligns with the low prevalence of hypertension among young people (Narchi, 2011; WHO, 2023b). A large portion of participants belonged to middle-aged adults between 40–65 years while older adults (65+) showed higher vulnerability to hypertension development. The age group of young adults between 18–40 years appeared less frequently in the studies, which conveys the importance of early monitoring and prevention strategies, because risk factors tend to emerge during these years (Lieb & Vasan, 2008). The broad age range in this review makes the findings more applicable and shows the need for prevention and management strategies that fit different age groups.

### Presentation of Results by Key Themes

To facilitate the interpretation of the diverse results, studies were grouped thematically according to their conceptual focus and methodological approaches. The results are presented under five main themes which correspond to different psychological dimensions that are associated with hypertension. Themes are as follows: (1) Psychological Disorders and Hypertension: Depression, Anxiety, and PTSD, (2) Chronic Stress, Emotional Reactivity, and Behavioural Risk Factors, (3) Psychosocial Interventions and Psychological Resilience in Hypertension Management, (4) Socioeconomic Influences on Hypertension–Psychological Links, (5) Vulnerable and Marginalised Populations

#### Theme 1 - Psychological Disorders and Hypertension: Depression, Anxiety, and PTSD

This theme draws from 60 studies which examine the relationship between depression, anxiety, PTSD and hypertension. A majority of 55 studies found moderate to strong positive correlations, 3 found inverse relationship, while 2 reported no association (see Table 1). For the strength of association between depression, anxiety and hypertension, nineteen studies found strong effects odds ratio (OR > 2.0), while thirty-six studies found moderate effects odds ratio (OR = 1.3 -- 2.0). These findings show psychological distress is a significant factor in hypertension risk.

**Table 1.**
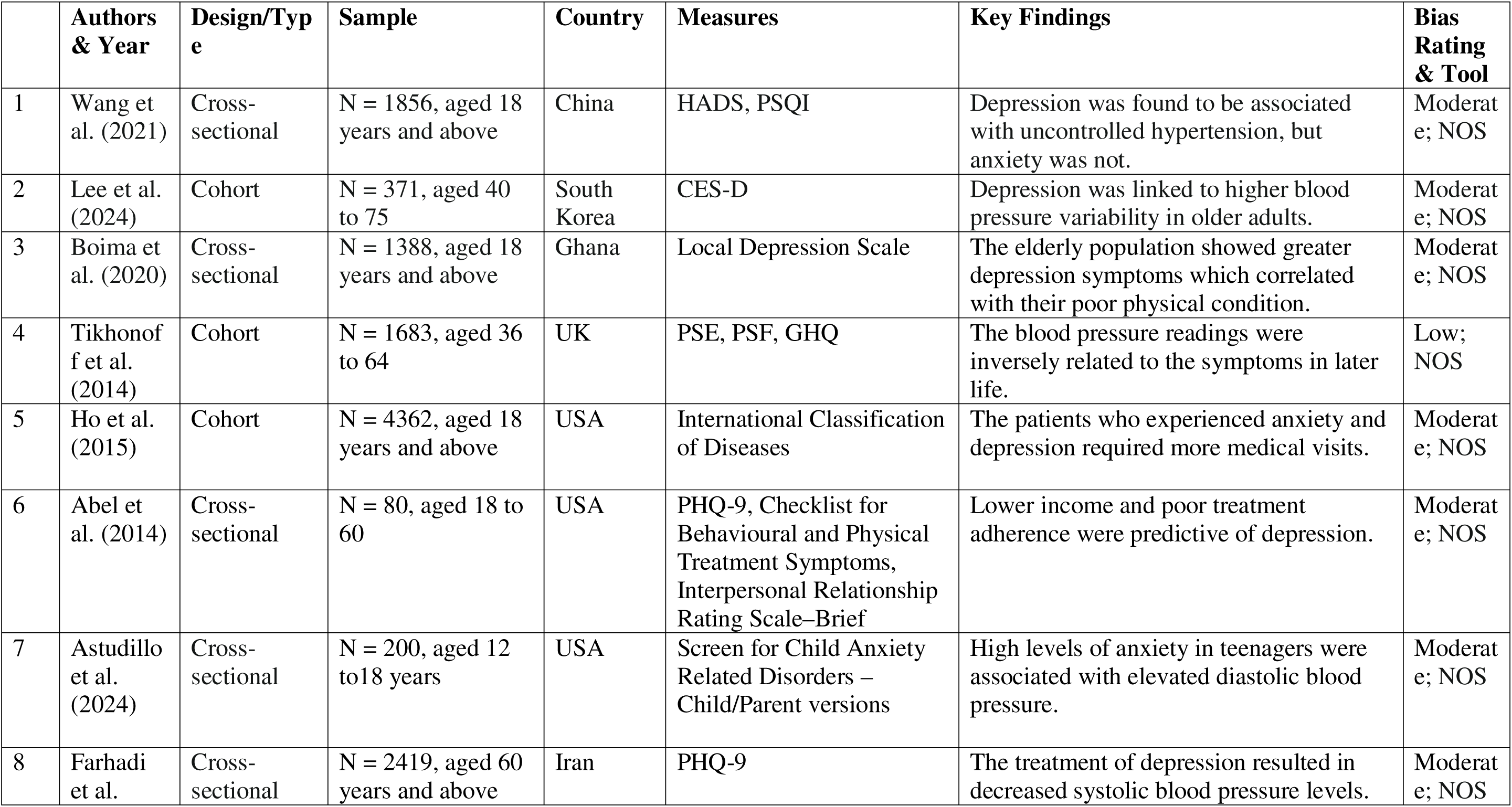
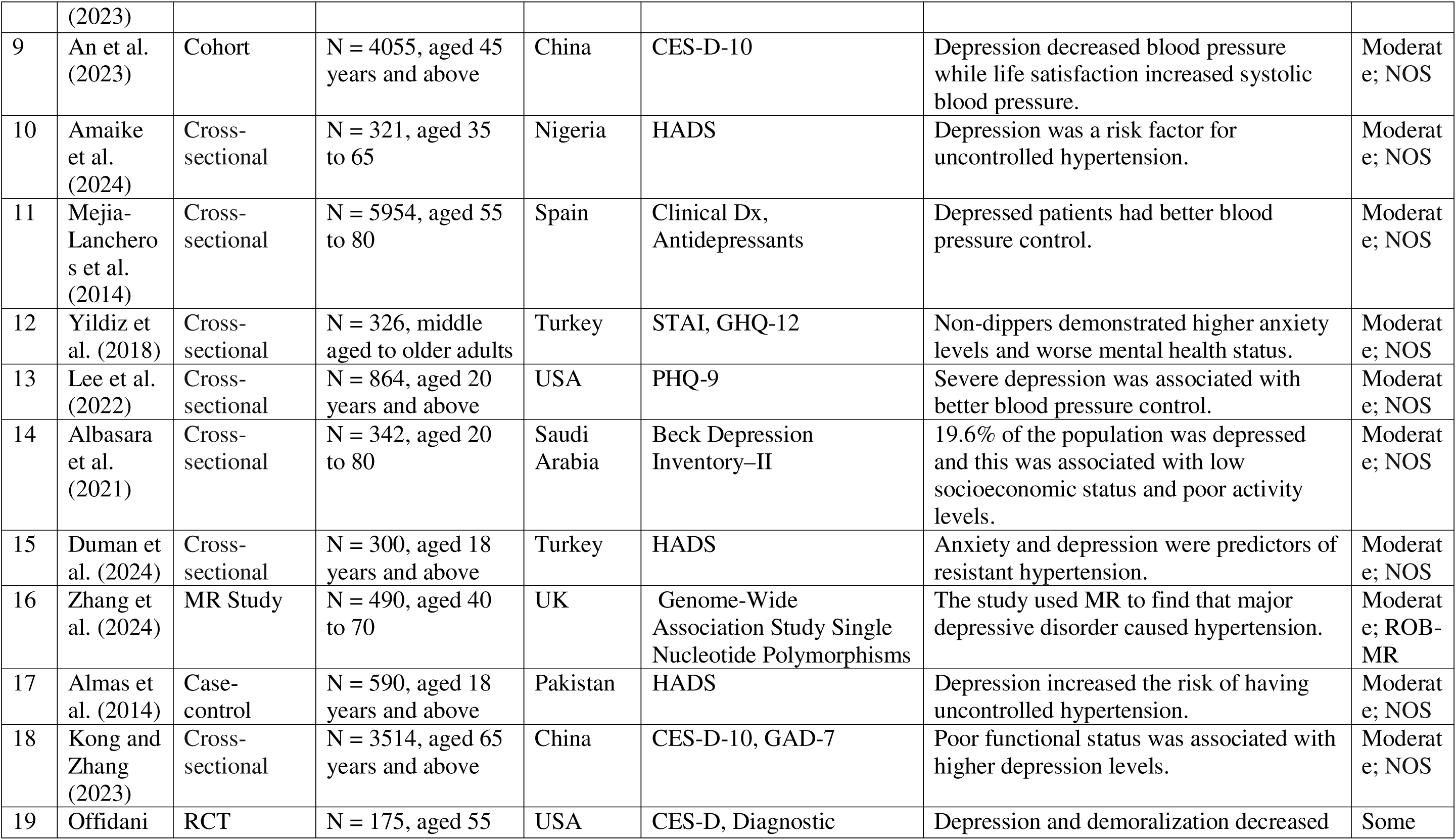
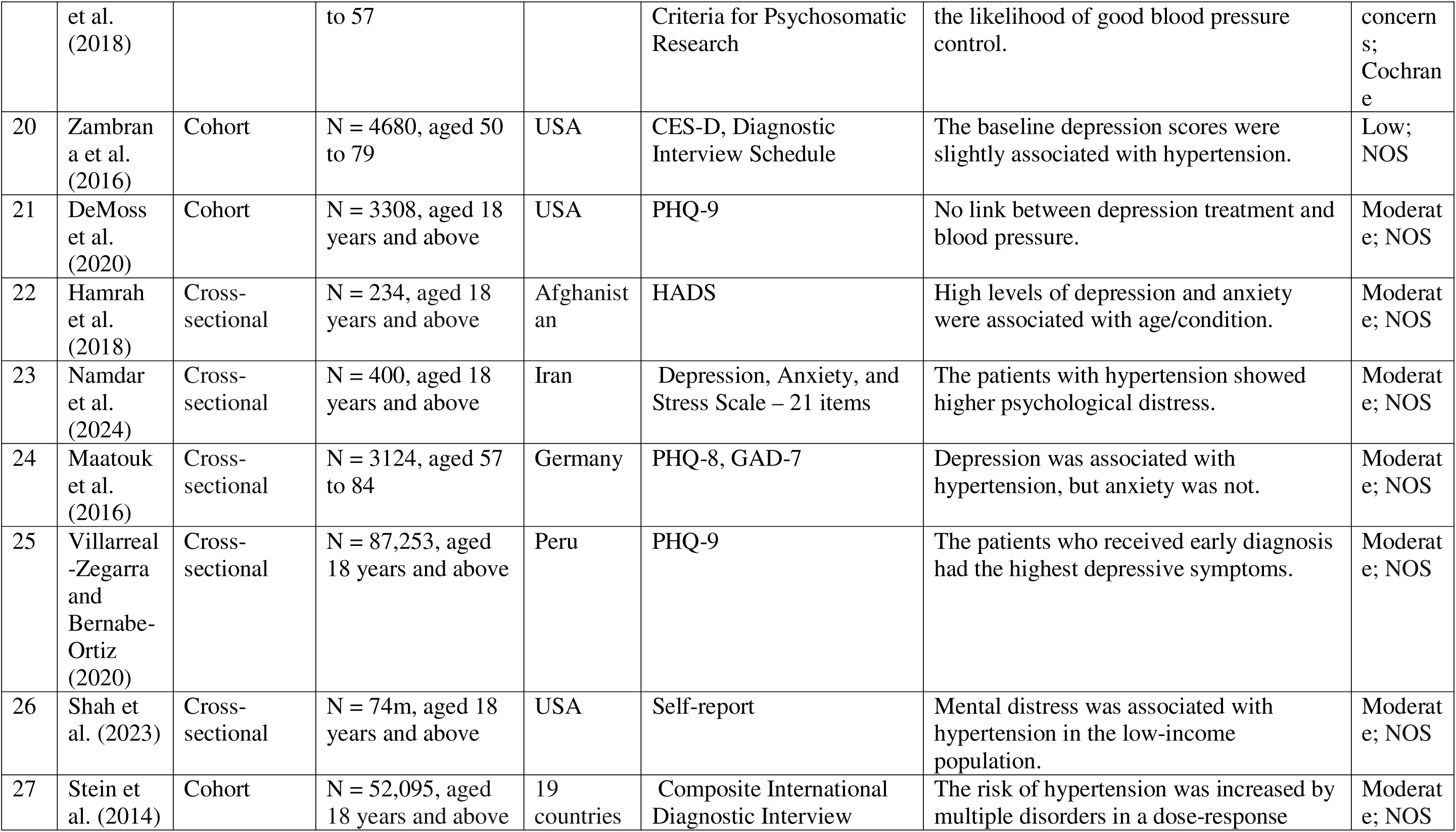
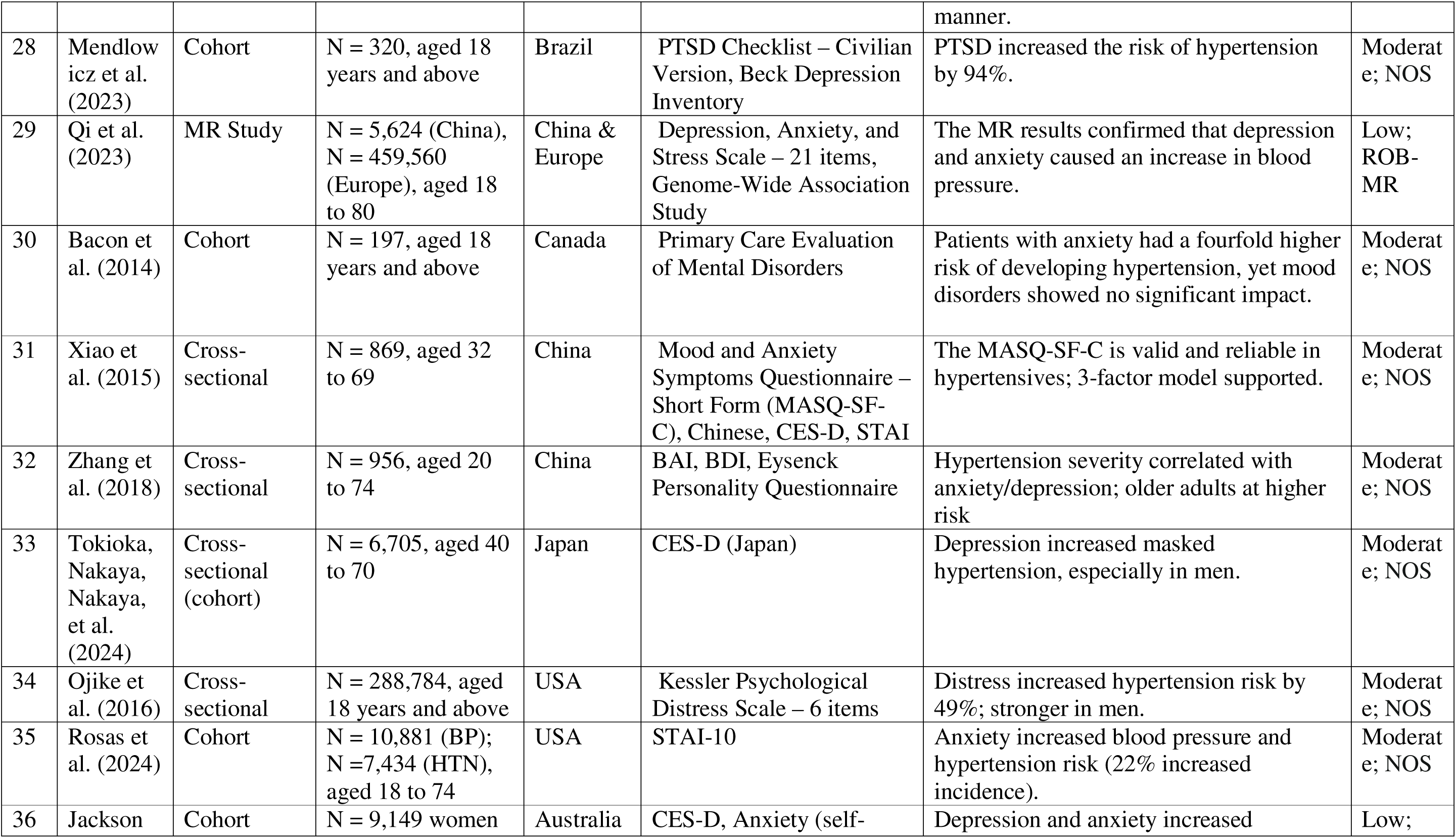
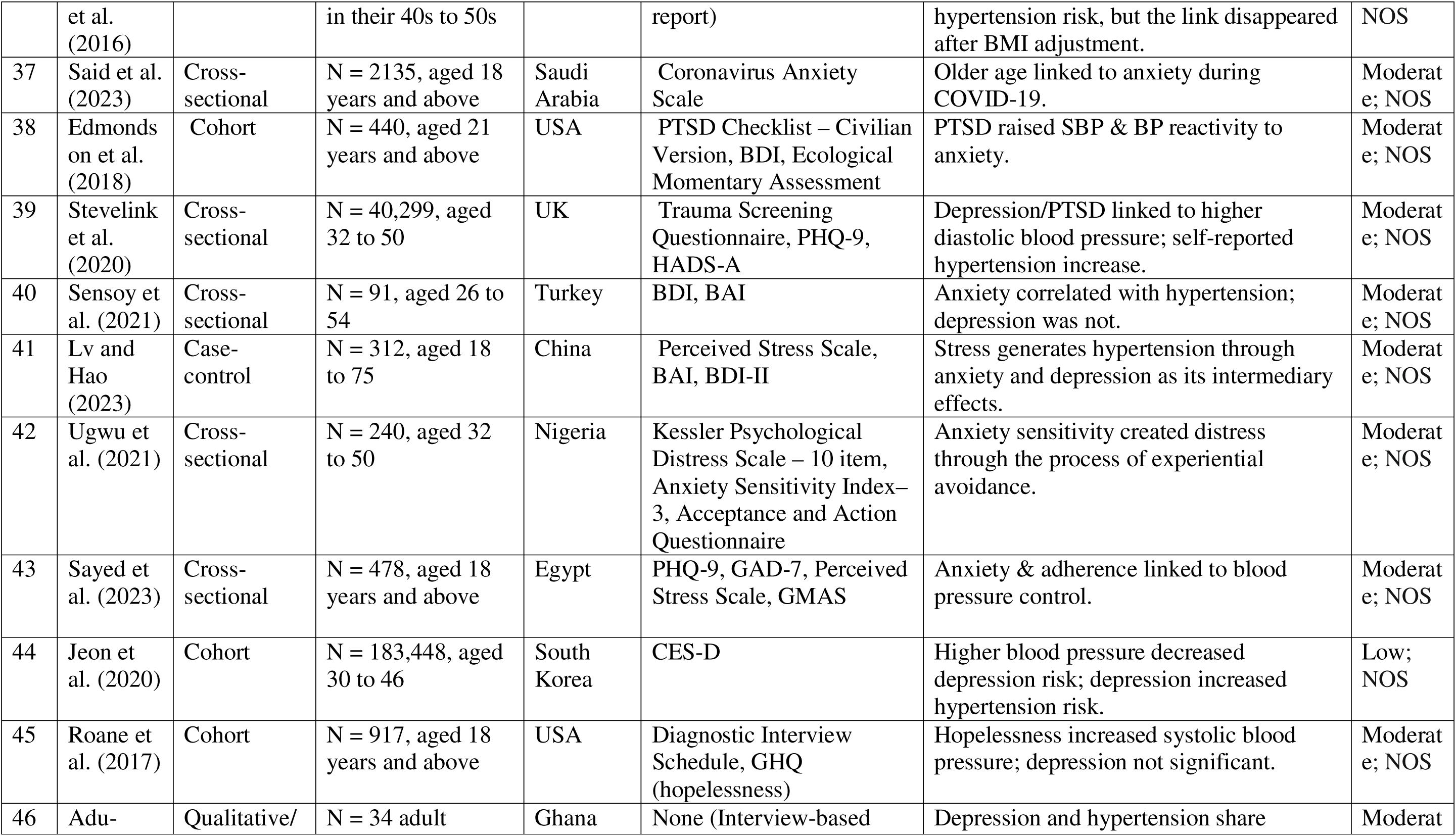
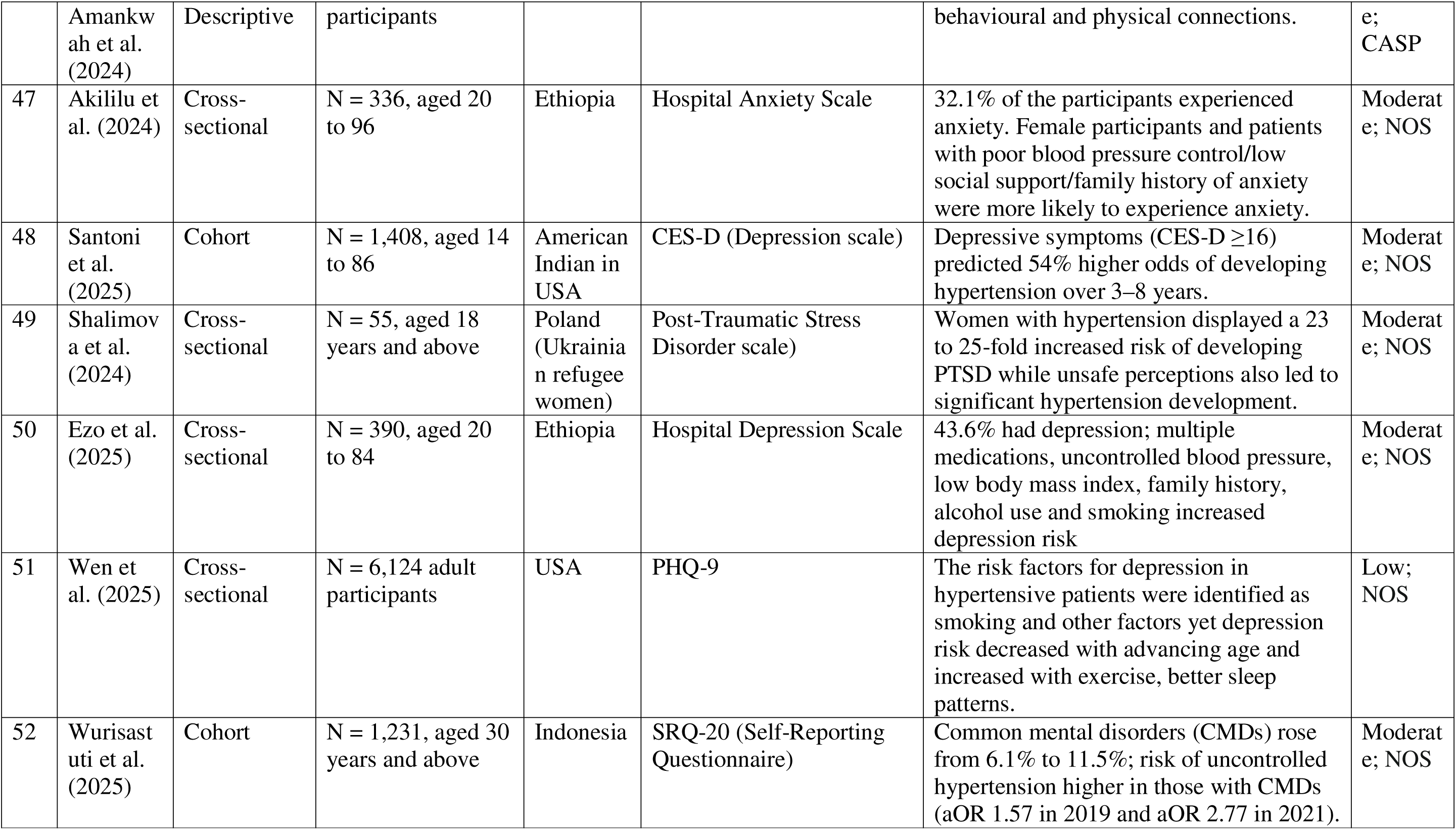
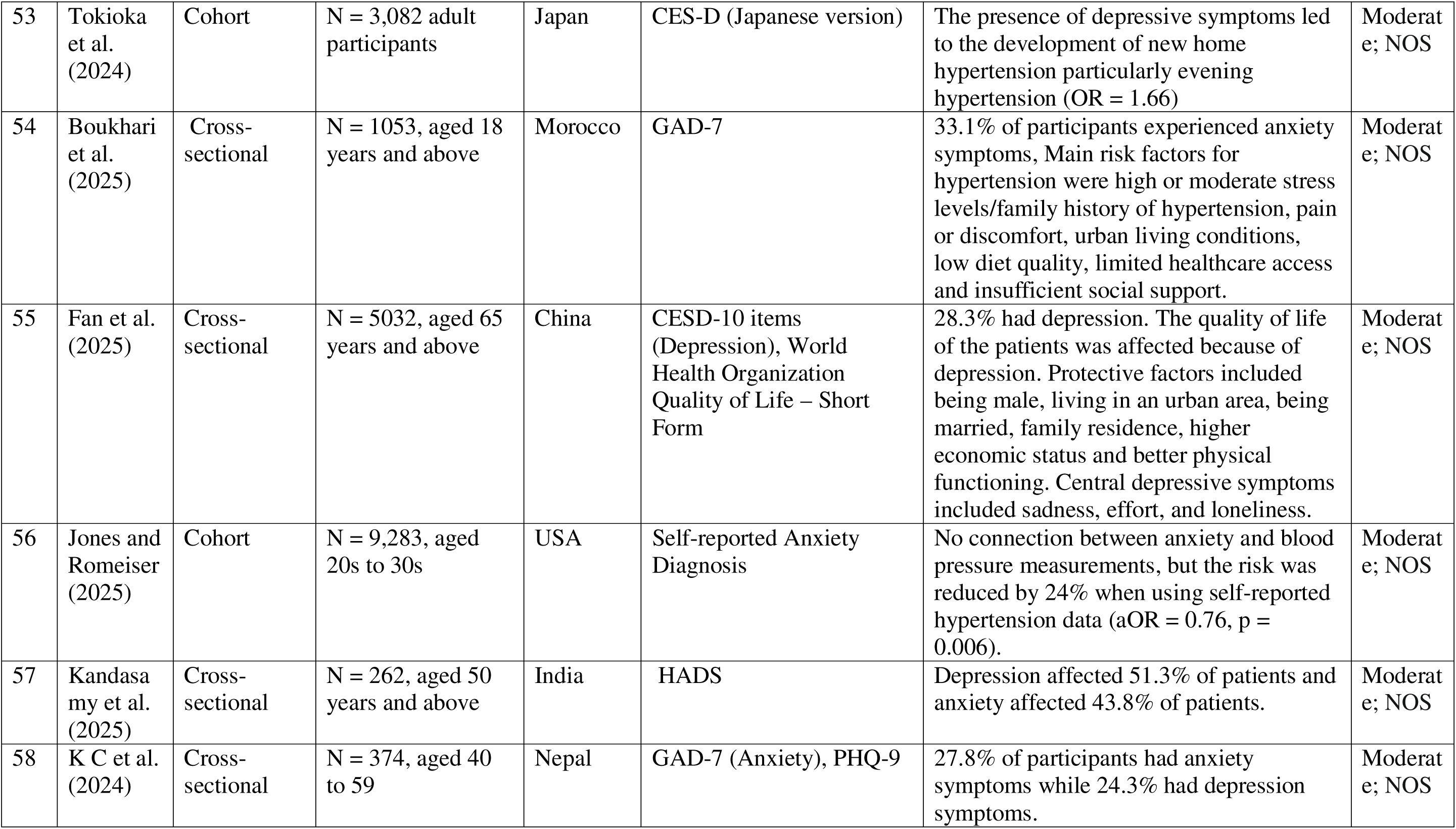
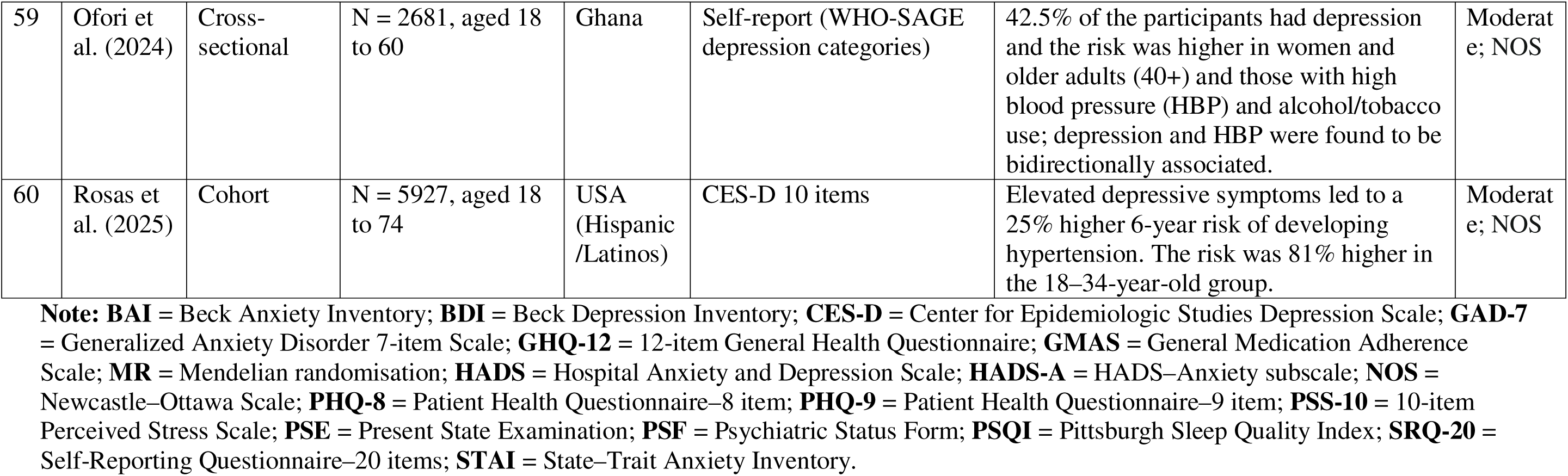
Theme 1: Psychological Disorders and Hypertension – Depression, Anxiety, and PTSD.

Research conducted by Stein et al. (2014) and Mendlowicz et al. (2023) established a significant connection between mental disorders and PTSD to increased hypertension risk especially when individuals face high-stress environments. Qi et al. (2023) used Mendelian Randomization to establish anxiety, depression and stress directly cause high BP. In Zhang et al. (2018) report, they conveyed that anxiety and depression were associated with higher hypertension levels.

Tokioka et al. (2024) investigated the relationship between depressive symptoms and masked hypertension which presents as normal clinic BP but elevated home BP. Their research showed that participants with depressive symptoms had a higher risk of developing masked hypertension particularly among male participants, which demonstrates that depression as a mental health condition increases the risk of developing undiagnosed hypertension. Ojike et al. (2016) conveyed that psychological distress such as how people felt (whether sad, worthless, hopeless, nervous) increased the risk of hypertension after controlling for other potential factors (such as age, sex, education level, race, income, marital status).

Bacon et al. (2014) longitudinal study notably demonstrated anxiety leads to a fourfold increase in hypertension risk for one year. Similarly, Rosas et al. (2024) found anxiety symptoms predicted changes in BP during six years and hypertension incidence. However, Akililu et al. (2024) report showed that anxiety is not an isolated condition, rather, the link with high BP is associated with how well individuals manage their BP, the gender they are in, and their social support system.

Further evidence by Adu-Amankwah et al. (2024) performed qualitative research which confirmed depression and hypertension are commonly believed to cause each other through emotional distress and behavioural changes while supporting integrated care approaches for both conditions. But not all studies aligned with this view. Evidence from the literature was mixed.

Two studies found no significant link, possibly due to differences in sample characteristics or research design (DeMoss et al., 2020; Zambrana et al., 2016). Other studies found inverse relationships such as depression influenced better BP control (Lee et al., 2022); anxiety and depression influenced lower systolic BP (Tikhonoff et al., 2014); while anxiety lowered the risk of developing hypertension in young and middle-aged adults (Jones & Romeiser, 2025), which provide preliminary evidence for the emotional dampening hypothesis theory.

#### Theme 2 - Chronic Stress, Emotional Reactivity, and Behavioural Risk Factors

This theme analysed stress-related elements which extend past clinical anxiety and depression. Among 28 studies, two found no association, two reported inverse association, 20 identified moderate positive associations between stress and elevated BP, and four studies revealed strong associations which confirmed chronic stress as a major hypertension risk factor (see Table 2).

**Table 2.** Theme 2: Chronic Stress, Emotional Reactivity, and Behavioural Risk Factors.

|  | Authors & Year | Design/Type | Sample | Country | Measures | Key Findings | Bias Rating & Tool |
| --- | --- | --- | --- | --- | --- | --- | --- |
| 1 | Hu et al. (2015) | Cross-sectional | N = 5976, aged 40 to 60 | China | Composite International Diagnostic Interview – Short Form | The relationship between stress and high blood pressure risk is stronger for women than men. | Moderate; NOS |
| 2 | Lu et al. (2019) | Cross-sectional | N = 530, aged 18 and above | USA | PSS, Duke Functional Social Support Scale, Social Network Index | Hypertension risk increases when stress levels rise yet social support networks lead to decreased hypertension risk. | Moderate; NOS |
| 3 | Schutte et al. | Cohort | N = 107, aged | South | Kessler Psychological | Distress led to hypertension | Moderate; |
|  | (2015) |  | 30 to 75 | Africa | Distress Scale – 6 items | development during the five-year period and confirmed alcohol consumption as an additional contributing factor. | NOS |
| 4 | Tomitani et al. (2022) | Cohort | N = 50, aged 50s to 70s | Japan | HeartGuide Wearable Blood Pressure Monitor + self-report | Stress and negative emotions caused blood pressure to rise by 15 mmHg. | Moderate; NOS |
| 5 | Gianaros et al. (2017) | Cross-sectional | N = 310, aged 30 to 51 | USA | fMRI, Stroop Colour and Word Test, Multi-Source Interference Task | The brain's activation patterns were able to forecast how blood pressure would change in response to stress. | Moderate; NOS |
| 6 | Ifeagwazi et al. (2018) | Cross-sectional | N = 226, aged 40 to late 60s | Nigeria | Emotional Reactivity Scale, State–Trait Anxiety Inventory | Anxiety acted as a connecting factor that produced both elevated blood pressure and emotional reactivity. | Moderate; NOS |
| 7 | Symonides et al. (2014) | Cross-sectional | N = 195, aged 30 to 60 | Poland | Courtauld Emotional Control Scale | The inability to control emotions resulted in poor blood pressure management. | Moderate; NOS |
| 8 | Jadhav et al. (2014) | Cross-sectional | N = 3600, aged 40 and above | India | Presumptive Stressful Life Events Scale | Mental stress proved to be a risk factor for hypertension exclusively in male participants. | Moderate; NOS |
| 9 | Mucci et al. (2016) | Cross-sectional | N = 412, aged 16 to 31 | Italy | GHQ-12 | Stress-related factors explained additional blood pressure variance yet only 3-4% of the explained variance was accounted for. | Moderate; NOS |
| 10 | Hassoun et al. (2015) | Cross-sectional | N = 3352, aged 18 to 75 | Germany | Trier Inventory for Chronic Stress – Screening Scale for Chronic Stress | blood pressure showed an inverse relationship with stress levels particularly among female participants. | Moderate; NOS |
| 11 | Li et al. (2023) | Cross-sectional | N = 10,823, aged 18 and above | China | PSS-14 items | A negative association exists between stress levels and hypertension risk. | Moderate; NOS |
| 12 | Spruill et al. (2019) | Cohort | N = 1,829 Black adults, aged 21 and above | USA | Global Perceived Stress Scale, Weekly Stress Inventory | High stress levels were associated with an increased risk of hypertension among women. | Low; NOS |
| 13 | Crump et al. (2016) | Cohort | N = 1,547,182 male conscripts, aged 18 | Sweden | Stress Resilience Scale | Individuals with lower resilience levels developed hypertension problems mostly when their body mass index was high. | Moderate; NOS |
| 14 | Roohafza et al. (2022) | Cohort | N = 3,428, aged 35 and above | Iran | GHQ-12, Stressful Life Events Questionnaire | Stress increases the risk of hypertension most notably among female participants. | Moderate; NOS |
| 15 | Dich et al. (2020) | Cohort | N = 7,479, aged 34 to 56 | UK | GHQ, Affect Scale | Emotional levels above average increased blood pressure measurements yet average emotional levels led to better blood pressure results. | Low; NOS |
| 16 | Trudel-Fitzgerald et al. (2014) | Cohort | N = 6,384, aged 39 to 63 | UK | Emotional Vitality Scale | Hypertension risk decreased when patients felt vital while optimism failed to show any significant relationship. | Low; NOS |
| 17 | Lakatos et al. (2022) | Case-control | N = 276, aged 40 to 65 | Romania | DECAS Personality Inventory | People with low emotional stability faced a 3.55 times higher risk of developing hypertension. | Moderate; NOS |
| 18 | Arroyave-Atehortua et al. (2023) | Cross-sectional | N = 79, aged 25 to 70 | Colombia | NEO Five-Factor Inventory, Ruminative Responses Scale | Perseverative cognition did not function as a mediator in this study, yet neuroticism maintained its connection to cognitive factors. | Moderate; NOS |
| 19 | Spartano et al. (2014) | Lab experimental | N = 25, aged 18 to 58 | USA | Tonometry, Doppler, Oscillometric | Carotid SBP reactivity had a direct impact on Intima–Media Thickness while brachial systolic blood pressure did not produce any significant findings. | Moderate; Domains approach |
| 20 | O'Connor et al. (2017) | Lab experimental | N = 90, aged 18 and above | UK | Cognitive Control Scale, Stressor Appraisal, Maastricht Acute Stress Test | High self-control produced two effects: it decreased blood pressure reactivity levels and enhanced blood pressure recovery. | Moderate; Domains approach |
| 21 | Kang et al. (2018) | Cross-sectional | N = 571 Black women, aged 18 to 60. | USA | Single-item stress scale | Higher stress levels among the participants resulted in elevated SBP readings and poor BP control due to non-pharmacological non-adherence. | Moderate; NOS |
| 22 | Stueck et al. (2016) | Cross-sectional | N = 79, aged 35 to 50 | Germany | AVEM-Work-related Behaviour and Experience Patterns Questionnaire | Patients with hypertension displayed higher stress levels than those with hypotension while the latter group exhibited worse anger control. | Moderate; NOS |
| 23 | Lin et al. (2020) | Cohort | N = 1112, aged 18 to 40. | Taiwan | Brief Symptom Rating Scale – 5 items | No significant connection between stress levels and blood pressure variability. | Low; NOS |
| 24 | Schaare et al. (2023) | Cohort | N = 502,494, aged 37 to 73. | UK | UK Biobank Mental Health Assessment Tools, fMRI | Hypertension was found to decrease both well-being and amygdala response. | Low; NOS |
| 25 | Tompson et al. (2019) | Pre-post | N = 183, aged 40 to 85 | UK | HADS, EuroQol 5-Dimension Scale | The procedure of out-of-office BP monitoring creates stress for certain patients although it does not result in clinical anxiety or depression. | Moderate; Domains approach |
| 26 | Jagtap et al. (2024) | Cross-sectional | N = 231 university employees, aged 18 | India | PSS-10 | People with moderate or high stress levels experienced 2.5 times more hypertension risks while 39% of the participants had hypertension and both age and gender proved significant factors. | Moderate; NOS |
| 27 | Aldirawi et al. (2024) | Cross-sectional | N = 276 male patients, aged 20 and above | Jordan | DASS-42 (Stress Subscale) | 55.1% of participants experienced psychological stress. Stress connected to chronic illness, inactivity, work/home stress, poor sleep and multiple social/clinical factors were predictors of stress. | Moderate; NOS |
| 28 | Balachandran et al. (2025) | Cross-sectional | N = 234 university teachers, aged 30 to 50. | India | Teacher Stress Inventory | 83.7% of participants had moderate-to-high stress levels and age $\leq 45$ combined with low physical activity was linked to stress and moderate stress increased hypertension risk by 2.78 times. | Moderate; NOS |

**Note:** **DASS-42** = Depression, Anxiety, and Stress Scale – 42 items; **fMRI** = Functional Magnetic Resonance Imaging; **GHQ** = General Health Questionnaire; **HADS** = Hospital Anxiety and Depression; **NOS** = Newcastle–Ottawa Scale; **PSS** = Perceived Stress Scale; Scale;

Brain activity due to stress explained 9% of BP fluctuations (Gianaros et al., 2017). Jagtap et al. (2024) discovered that university staff members who experienced moderate to high stress levels had a 2.5 times greater chance of developing hypertension. Further insights revealed teachers along with other workplace employees developed hypertension because of their high levels of emotional strain which led to elevated BP (Stueck et al., 2016). Longitudinal studies revealed moderate but significant associations including Crump et al. (2016) and Roohafza et al. (2022) which demonstrated psychological factors (e.g., stress, mental health states) influencing long term hypertension. The research by Dich et al. (2020) revealed that both low and high negative emotions create increased BP risk while Trudel-Fitzgerald et al. (2014) discovered that emotional vitality (feeling energetic and engaged with life) acted as protection, but optimism did not reduce risk.

Spruill et al. (2019) discovered chronic stress elevates hypertension risk specifically among Black adults. The research by Ifeagwazi et al. (2018) and Sayed et al. (2023) convey psychological factors such as emotional reactivity contribute to elevated BP and poor hypertension management thus requiring mental health support in hypertension care particularly in LMICs. Kang et al. (2018) study revealed Black women who failed to control their stress levels followed hypertension management lifestyle recommendations (such as dietary changes, physical activity, limiting alcohol consumption, and weight loss) poorly which resulted in elevated BP and inadequate BP management.

Other reports showed lifestyle elements and psychosocial connections including physical activity help regulate stress-induced hypertension especially in young people (Mucci et al., 2016). Further evidence showed that personality emerged as a key factor. The research conducted by (Lakatos et al., 2022) demonstrated that lower emotional stability (high neuroticism levels) in humans represents major risk factors for developing hypertension. The research conducted by physiological studies supports these discovered connections. For example, Spartano et al. (2014) established that carotid BP (neck artery pressure) reactions to mental stress are linked to vascular risk factors while O’Connor et al. (2017) discovered self-control functions as a protective mechanism against BP changes, thus showing how cognitive and emotional traits influence hypertension risk. Other studies found that there was no significant association between stress and BP variability (Arroyave-Atehortua et al., 2023; Lin et al., 2020). Two studies (Hassoun et al., 2015; Li et al., 2023) reported inverse associations which support the emotional dampening hypothesis.

#### Theme 3 - Psychosocial Interventions and Psychological Resilience in Hypertension Management

Among the six intervention studies that utilized psychological and behavioural strategies to enhance hypertension outcomes, 5 had strong associations and 1 moderate. Overall, interventions which focus on psychosocial and psychological resilience in hypertension, produce positive effects on BP and psychological health.

**Table 3.** Theme 3 - Psychosocial Interventions and Psychological Resilience in Hypertension Management.

|  | **Authors & Year** | **Design/Type** | **Sample** | **Country** | **Measures** | **Key Findings** | **Bias Rating & Tool** |
| --- | --- | --- | --- | --- | --- | --- | --- |
| 1 | Hernandez et al. (2019) | RCT | N = 126 Hispanic patients, aged 18 and above | USA | Center for Epidemiologic Studies Depression Scale, Perceived Stress Scale, Fitbit | Positive intervention improved blood pressure and psychological well-being | Low; Cochrane |
| 2 | Hernandez et al. (2020) | RCT | N = 70 Hispanic patients aged, 18 and above | USA | Center for Epidemiologic Studies Depression Scale, PSS, Diabetes Empowerment Scale | Web-based intervention improved blood pressure and emotional health | Low; Cochrane |
| 3 | Momeni et al. (2016) | RCT | N = 60 cardiac patients, aged 40 to 55 | Iran | Perceived Stress Scale – 14 items, State–Trait Anger Expression Inventory–2 | Mindfulness-based stress reduction lowered systolic blood pressure, stress, anger; no Diastolic Blood Pressure effect | Moderate; Cochrane |
| 4 | Still and Ruksakulpiwat (2024) | Cross-sectional | N = 30 African Americans, aged 25 | USA | Perceived Stress Scale – 10 items, Patient-Reported Outcomes Measurement Information System tools | Resilience linked to lower stress, better health-related quality of life and disease self-efficacy | Moderate; Newcastle–Ottawa Scale |
| 5 | M. Lee et al. (2024) | RCT | N = 100 hypertensive patients, aged 60 and above | Taiwan | World Health Organization – Five Well-Being Index | Mobile app intervention improved mental health and self-management significantly vs. control. Effects seen in both main effect and interaction with time. Significant improvements post 3-month intervention. | Low; Cochrane |
| 6 | Parlak and Şahin (2025) | Pre-post intervention | N = 24 elderly hypertensive patients, aged 65 and above | Turkey | Geriatric Depression Scale, Perceived Stress Scale | 12-week creative handicraft intervention program significantly reduced stress and depression and helped regulate blood pressure; stress reduction linked to lower depression levels | Moderate; Domains approach |

An 8 week culturally tailored positive psychology intervention together with its digital version achieved substantial BP improvements in outcomes for Hispanic/Latino participants (Hernandez et al., 2020; Hernandez et al., 2019). MBSR intervention practice (such as yoga, gentle breathing, body scan) led to decreases in patients systolic BP together with reduced perceived stress and anger among cardiac patients (Momeni et al., 2016).

The research by (Lee et al., 2024) showed that mobile app-based self-management programs successfully improved mental health and self-management capabilities in patients with hypertension. The Mobile Application Disease Self-Management Programme (MADSMP) delivered educational content and behavioural support to hypertensive patients through a mobile application. The program educated patients about hypertension while teaching them to follow their medication and maintain healthy eating habits and daily lifestyle reminders for better self-care management of their condition. A 12-week creative handicrafts program developed by Parlak and Şahin (2025) successfully decreased depression, stress and lowered BP in elderly patients with hypertension. Further findings demonstrated that resilience framework together with self-efficacy play crucial roles in patient outcomes. African American individuals who demonstrated higher resilience experienced lower stress levels and practiced better self-care, highlighting psychological wellbeing improves hypertension outcomes (Still & Ruksakulpiwat, 2024).

#### Theme 4 - Socioeconomic Influences on Hypertension–Psychological Links

Six studies demonstrated both general and context specific patterns in their findings between socioeconomic factors and hypertension, revealing varying degrees of association. 2 had strong association, 3 moderate, 1 low (see Table 4).

**Table 4.** Theme 4 - Socioeconomic Influences on Hypertension–Psychological Links.

|  | **Authors & Year** | **Design/Type** | **Sample** | **Country** | **Measures** | **Key Findings** | **Bias Rating & Tool** |
| --- | --- | --- | --- | --- | --- | --- | --- |
| 1 | Kang & Kim | Cross-sectional | N = 29,425, aged 20 and above | South Korea | EuroQol 5-Dimension Scale, self-reported | The link between hypertension and depression depends on income level | Low; Newcastle– |
|  | (2020) |  |  |  | depression | because the high-income group shows a positive relationship, but the low-income group shows an inverse association. | Ottawa Scale |
| 2 | Vallée et al. (2021) | Cross-sectional | N = 66,478, aged 35 to 61 | France | Center for Epidemiologic Studies Depression Scale | Higher systolic blood pressure (SBP) levels were found in people who experienced depression, but socioeconomic status influenced the relationship between depression and SBP differently based on education level especially among women. | Low; Newcastle–Ottawa Scale |
| 3 | Luo et al. (2023) | Cohort | N = 11,050, aged ≥45 and above | China | Center for Epidemiologic Studies Depression Scale – 10 items | Obesity in combination with depression elevated the risk of developing hypertension and this combined effect explained 67% of total hypertension cases. | Low; Newcastle–Ottawa Scale |
| 4 | Wang et al. (2023) | Cross-sectional | N = 9,900, aged 40 to 70 | China | Patient Health Questionnaire–9 item, Generalized Anxiety Disorder 7-item Scale | The combination of hypertension and depression/anxiety symptoms mostly affected women who belonged to lower socioeconomic status. | Moderate; Newcastle–Ottawa Scale |
| 5 | Mushtaq and Najam (2014) | Cross-sectional | N = 237, aged 30 to 65 | Pakistan | Depression, Anxiety, and Stress Scale | The combination of depression with anxiety and stress produced hypertension symptoms which also proved predictive based on gender and social context. | Moderate; Newcastle–Ottawa Scale |
| 6 | Forsyth et al. (2014) | RCT | N = 463, aged 18 and above | USA | Scale of Racial and Ethnic Discrimination, Perceived Stress Scale, Patient Health Questionnaire–9 item, Morisky | Discrimination lowered medication adherence; depression was a key mediator. | Some concerns; Cochrane |

Older Chinese adults with low socioeconomic status (SES) who had depression and obesity showed increased risk for developing hypertension (Luo et al., 2023; Wang et al., 2023). Studies in South Korea and Pakistan revealed that SES outcomes between psychological health and hypertension are primarily influenced by income differences, gender expectations and mental health stressors (Kang & Kim, 2020; Mushtaq & Najam, 2014). French women with higher SES managed to keep their BP under control even when they experienced depression (Vallée et al., 2021). In the United States, discrimination perceptions lead to worse medication adherence because it creates more psychological distress (Forsyth et al., 2014). Overall, the findings highlight the need for interventions which use cultural perspectives to address socioeconomic conditions in order to enhance hypertension care through structural inequality reduction.

#### Theme 5 - Vulnerable and Marginalised Populations

This theme investigates depression as an independent outcome among marginalised hypertensive patients across different regions including Ghana and Nigeria (Ademola et al., 2019), Indonesia (Agustiya et al., 2024), Ethiopia (Abate et al., 2024), U.S. (Spikes et al., 2020), China (Li et al., 2025) and the Netherlands (Fernald et al., 2021) which demonstrates how cultural, social and structural elements affect prevalence and risk (see Table 5).

**Table 5.**
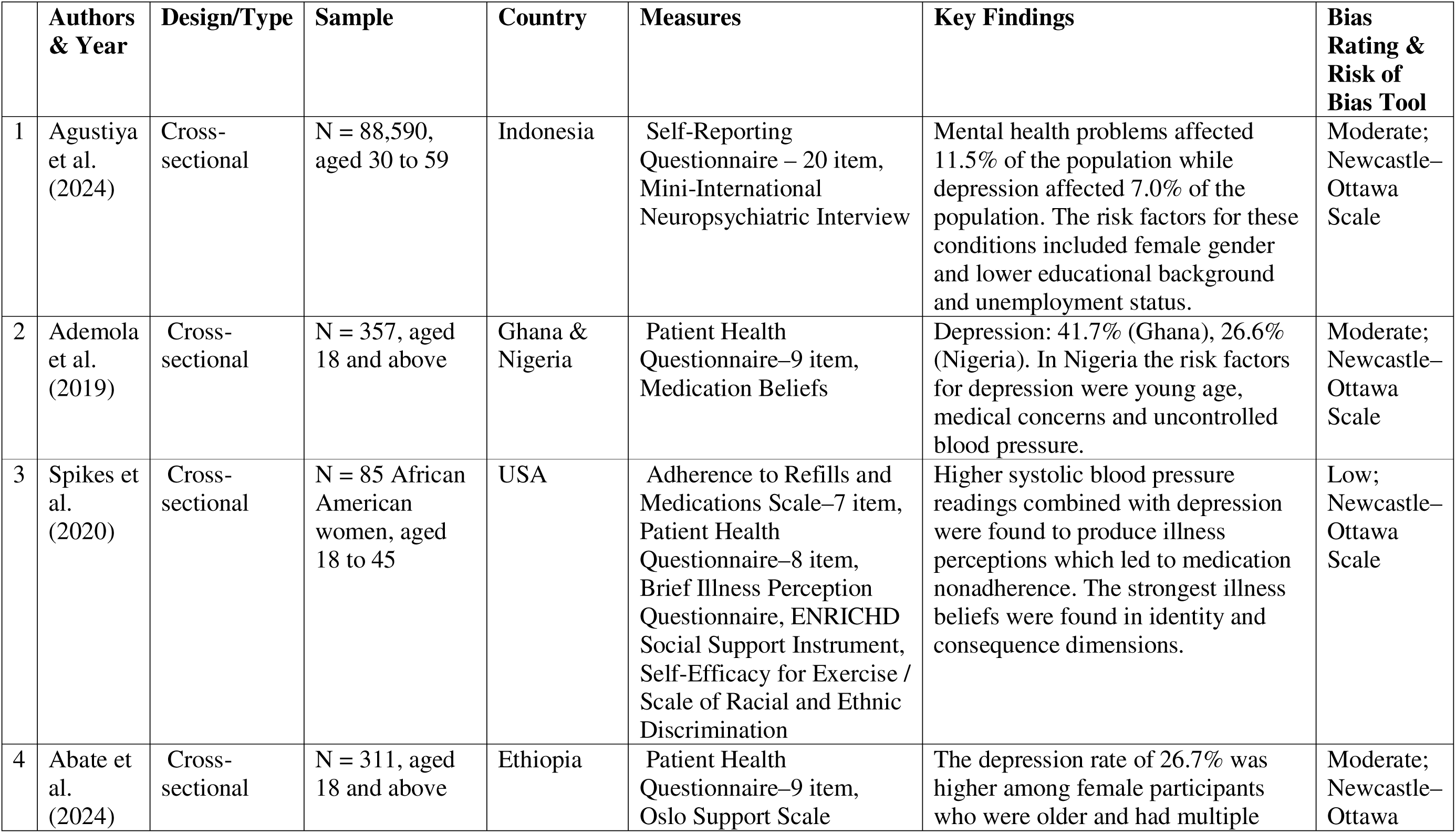
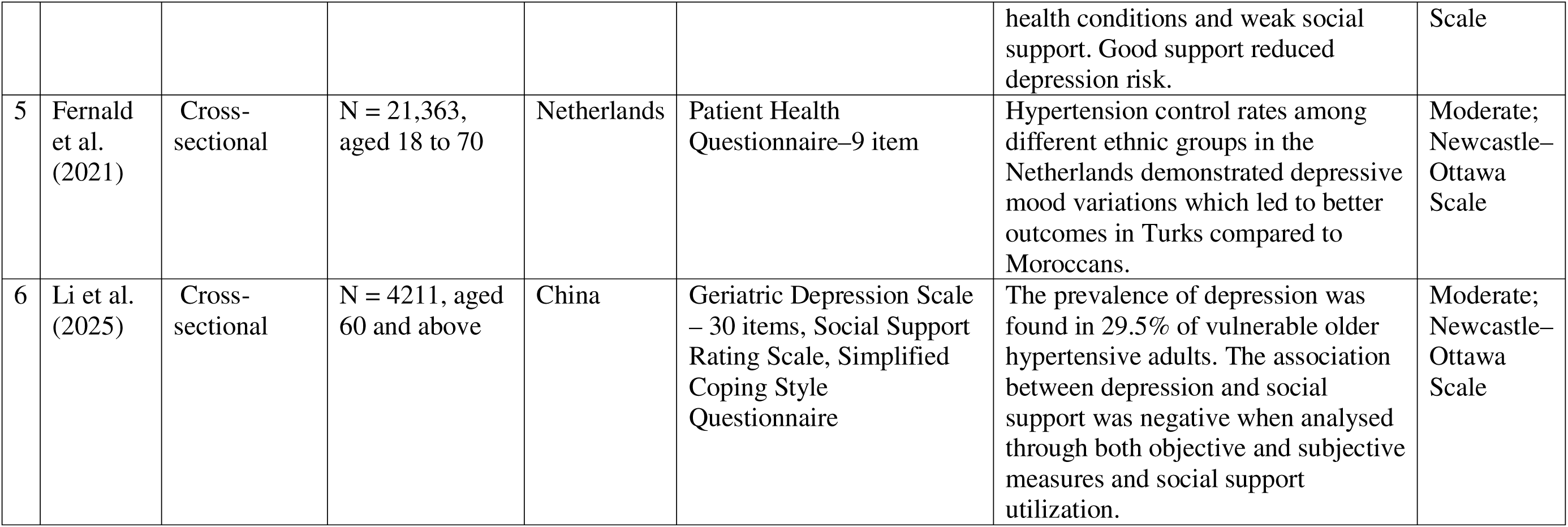
Theme 5 - Vulnerable and Marginalised Populations.

Depression rates showed substantial variations between the studied populations with Ghana at 41.7%, Ethiopia at 26.7%, Netherlands at 29.4% for Turkish and 26.4% for Moroccans, Nigeria at 26.6%, the U.S. at 24% and Indonesia at 7.0%. The observed differences between these populations stem from unequal healthcare availability, social discrimination and health-related social determinants.

The main risk factors were low educational attainment and joblessness in Indonesia. Nigerian patients were younger and had poor medication compliance, and Ethiopian older women had minimal social support. In the U.S., African American women experienced racial and gender specific stressors with 24% facing moderate depressive symptoms and more than 80% experiencing nonadherence to medication.

The HELIUS study (healthy life in an urban setting) revealed Turkish and Moroccan participants with significant depressed mood (SDM) experienced hypertension-related challenges because of their educational level and gender and socioeconomic status indicators (Fernald et al., 2021). Turkish participants achieved better BP control during their experience of SDM than Moroccan participants did. Li et al. (2025) found that depression occurred in 29.5% of older adults who had hypertension. Lower social support combined with negative coping styles led to higher depression levels.

## Discussion

This systematic review examined the complex link between psychological elements and hypertension through synthesizing findings from 2014 to 2025. The results demonstrate that psychological health plays a major role in both hypertension development and treatment management. Depression, anxiety, chronic stress and psychosocial factors act as primary psychological factors which increase BP and worsen hypertension control. Conversely, the study revealed that psychological resilience (Still & Ruksakulpiwat, 2024) together with mindfulness-based interventions (Hernandez et al., 2020; Hernandez et al., 2019) and strong social support networks (Spruill et al., 2019) serve as protective factors which help control hypertension while lowering BP levels.

The review demonstrates the significant role of how psychological disorders including depression and anxiety increase the risk of hypertension development. Mental health disorders including PTSD play a role in hypertension development especially when people work in high-stress environments (Amaike et al., 2024; Stein et al., 2014). The biopsychosocial model receives support from chronic stress research which shows that workplace stress intensifies hypertension while prolonged stress causes cardiovascular strain (Jagtap et al., 2024; McEwen, 2017). However, some studies align with the emotional dampening hypothesis that suggest stress may decrease blood pressure (Jones & Romeiser, 2025; Tikhonoff et al., 2014) although these results remain debated.

The effectiveness of MBSR as a psychosocial intervention has been demonstrated through its ability to manage hypertension while improving both BP and psychological health (Hernandez et al., 2019; Momeni et al., 2016). Mobile self-management programs have also shown their value in practice (Lee et al., 2024). The combination of low SES with psychological stress increases hypertension risk thus requiring additional research in LMICs (Luo et al., 2023; Vallée et al., 2021).

This review further conveys marginalized groups including those from LMICs and minority populations in high-income nations experience discrimination and restricted healthcare access which results in worse hypertension outcomes (Fernald et al., 2021; Spikes et al., 2020).

These findings demonstrate why culturally appropriate integrated healthcare systems must address both physical and mental health requirements.

### Study Strength and Limitation

The study’s strength is in its comprehensive literature review, identification of key psychosocial factors that influence hypertension, and the need for integrated care models that address both mental and physical health. However, there are limitations. This review identifies two main gaps in the literature which consist of insufficient qualitative research about hypertension patients’ personal experiences and the geographic focus on high-income countries. Qualitative research would help us better understand the complex relationships between psychological factors, cultural, social and environmental factors that affect hypertension management (Londoño Agudelo et al., 2021). Additionally, the findings lack generalizability because LMICs remain underrepresented in the literature, with most contributing to only one or two studies, but no more than four studies on the psychological impact of hypertension. This highlights the need for additional studies from sub-Saharan Africa, South Asia and Latin America where hypertension and mental health problems are prevalent due to cultural narratives which impact health behaviour (Renjith et al., 2021).

Moreover, the review emphasizes the necessity of adding psychological care to hypertension treatment protocols. The current focus on biological aspects and lifestyle modifications in treatment should be expanded by incorporating mental health interventions because research shows their effectiveness in improving results. Future research needs to investigate how cognitive-behavioural therapy, stress management, and psychosocial support can be incorporated into treatment plans for patients who have psychological comorbidities.

## Conclusion

Overall, this review provides evidence that the development and management of hypertension strongly depends on psychological factors which include depression alongside anxiety and stress. Mental health interventions demonstrate potential benefits, but additional research is required to create treatment models which address both psychological and physical aspects of hypertension. Research should focus on using qualitative approaches while studying diverse populations including those from LMICs to evaluate the extended impacts of psychosocial interventions. The management of hypertension requires an integrated approach which treats mental health with equal importance as physical health.

## Data Availability

All data produced in the present work are contained in the manuscript

