## Supplementary Materials for "Understanding the Psychological Factors that Impact Hypertension: A Systematic Review"

### **Search strategy**

| **Database** | **Search Strategy** | **Filters** |
| --- | --- | --- |
| PubMed, MEDLINE (via EBSCO), Embase, PsycINFO, Scopus, CINAHL | #1: ("hypertension"[MeSH] OR "blood pressure"[MeSH] OR hypertension*[tiab] OR "high blood pressure"[tiab])  #2: ("psychology"[MeSH] OR "psychological factors"[tiab] OR "mental health"[tiab] OR "anxiety"[tiab] OR "depression"[tiab] OR "stress"[tiab] OR "cognitive factors"[tiab] OR "worry"[tiab] OR "personality traits"[tiab] OR "psychological disorders"[tiab])  #3: #1 AND #2  #4: (hypertension* OR "blood pressure*") AND ("mental disorders"[tiab] OR "mental illness"[tiab] OR mind*[tiab])  #5: hypertension* AND "mental health"[tiab] | Published from 2014 onwards; English language; Humans; Peer-reviewed articles, clinical trials, systematic reviews, meta-analyses, RCTs, and reviews. |

**Key Notes on Search Strategy:**
 **#tags** are used to clearly indicate each stage of the search process within each database, facilitating ease of replication and transparency.
 **Truncations (**)*** *have been applied to capture multiple forms of words (e.g., "hypertensi**" retrieves hypertension, hypertensive, etc.).
 **Field-specific searches** are included (e.g., ti,ab for title and abstract fields) to enhance the precision of search terms.
 **Mesh Terms and Expansions** were used where available to ensure comprehensive retrieval of all relevant studies in the respective databases.
